# Feasibility and effectiveness of a smartwatch-based intervention program for ameliorating depressive symptoms: a pilot-study

**DOI:** 10.1101/2025.01.18.25320669

**Authors:** Jeanette Tamm, Andy Brendler, Anastasia Bauer, Janina Gordon, Daniela Strauss, Ronja Brinkmann, Raphael Degmayr, Daniel Pfahl, Markus Friedrichs, Julia Schwind, Victor Spoormaker

## Abstract

**Objective:** Depression remains one of the most critical healthcare concerns worldwide. While increasing patient numbers inevitably lead to a greater need for treatment, resources to adequately match this demand are limited. Digital technologies hold promises to complement standard therapeutic approaches such as psychotherapy, thereby alleviating the strain on an increasingly burdened healthcare system. This study aimed at testing a new comprehensive smartwatch-based digital intervention program administered for 12 weeks in depressed patients to aid therapeutic progression and symptom relief.

**Methods:** Seventeen mildly to moderately depressed patients receiving the smartwatch-based intervention were compared to an existing, retrospective control group (35 patients). The intervention program comprised behavioral activation, cognitive-behavioral intervention for insomnia and physical activity interventions. Intervention effects on the reduction of depression severity, measured by the PHQ-9, were investigated using a repeated measure analysis of variance (rmANOVA).

**Results:** The rmANOVA showed a significant group x time interaction for the PHQ-9 scores, *F*(1, 50) = 7.21, *p* = 0.010, with post-hoc t-tests revealing group differences after but not before the intervention.

**Conclusion:** This study suggests that smartwatch-based interventions are feasible and effective in reducing mild to moderate depressive symptoms and could therefore be a promising stand-alone or complementary intervention to psychotherapy. Further research with a randomized controlled trial is required to validate these results.

## Introduction

Depression is among the largest contributors to non-fatal disease burden and health loss worldwide (Patel et al., 2016) with an estimated annual medical and economic costs of more than €190 billion in the U.S. alone (Greenberg et al., 2023). These trends pose challenges for individual patients and the healthcare system as a whole, given the resulting high demand for treatment. Most commonly prescribed evidence-based treatments are psychopharmacological medication and psychotherapy. Among the latter, cognitive behavioral therapy (CBT) and in particular behavioral activation has been reported to have the strongest effects to mitigate mild to moderate depression (Cuijpers et al., 2020; Derubeis et al., 1999; Furukawa et al., 2021). Even though psychotherapy constitutes one of the most effective therapeutic approaches to date (Cuijpers et al., 2014), its largest problem lies in the high number of required resources to ensure adequate and efficient treatment (e.g., trained personnel, treatment facilities, and treatment time).

However, a promising solution addressing the discrepancy between the high therapeutic need and low availability of resources in the form of trained psychologists and psychotherapists are digital solutions. For example, digital CBT, i.e., administering therapy through the internet has been shown to be effective in numerous studies (Etzelmueller et al., 2020; Kambeitz-Ilankovic et al., 2022), albeit with lower effect sizes than face-to-face therapy. Furukawa et al. (2021) conducted a meta-analysis of 76 randomized controlled trials on digital interventions for major depressive disorder (MDD), and found evidence for web- or app-based behavioral activation being effective in ameliorating MDD, with weaker evidence suggesting the same for digital CBT for insomnia (CBT-I). These components can be effectively delivered through web-based and smartphone applications, given that they comprise questionnaires (e.g., lists of pleasurable activities), psychoeducational texts, subjective feedback inputs (e.g., recording bedtimes, sleep, and waking times) and activity scheduling in an agenda. Even though there are now various digital interventions that guide patients to more functional behaviors and cognitions on a weekly basis, low adherence rates (Kernebeck et al. 2021) and lack of monitoring of disease progression (Polhemus et al., 2022) remain key challenges.

In addition to behavioral activation and CBT-I, specific interventions targeting increased physical activity also reduce depressive symptoms. A recent meta-analysis of multiple meta-analyses (Singh et al., 2023) reported moderate effect sizes for symptom improvement in MDD. In order to incorporate physical activity interventions digitally, it would be helpful to objectively monitor and provide feedback on activity levels and behavior. To this end, applications are needed that go beyond classic web- and smartphone-based apps by incorporating data of sufficient quality from wearable devices such as smartwatches or fitness trackers.

A few previous studies incorporating smartwatches in relation to depression have shown promising trends in reducing symptoms (Aguilar-Latorre et al., 2022), however, the number of such studies is still limited and it has been suggested that more research in this direction is needed to successfully incorporate this technology in clinical practice (Triantafyllidis et al., 2024), for instance with feedback derived from smartwatches that is automatically integrated into the digital intervention.

To fill this gap, this study combines these beneficial aspects of digital interventions in a smartwatch-based intervention program. First, we incorporated more active physical activities, such as sports, into the behavioral activation component, resulting in three activity categories: social, relaxing, and sports activities. Second, we provided feedback on actual activity levels through step counts. Third, we used the smartwatch for ecological momentary assessments (EMA; e.g., subjective impression of sleep quality or depressed mood). Our intervention incorporated data from smartwatches, enabling the integration of behavioral data into the patient’s progress report and the presentation of recommendations for evidence-based actions. In this initial pilot-study (n=17), we aimed to evaluate the feasibility and initial effect sizes of this smartwatch-based intervention for patients with mild and moderate depression on the Patient Health Questionnaire (PHQ-9), and compared these initial effects to those of an existing, retrospective control group.

## Methods

### Participants and Recruitment

Participants were recruited in the framework of the Smartwatch-based Momentary Assessment and Real Time Technology (SMART) study, which was conducted at the Max Planck Institute of Psychiatry in Munich, Germany. The SMART study was a monocentric, single-arm digital intervention study with 12 weekly intervention modules for participants experiencing depressive symptoms. The primary outcome was the repeated assessment of the PHQ-9 (before and after the 12-week intervention). The study protocol was approved by the Institutional Review Board (“Ethics Committee”) of the Faculty of Medicine at LMU Munich (Project number 18–553). All participants provided written informed consent prior to clinical interviews and study participation.

Participants were recruited through online media announcements (e.g., Facebook, the institute’s website). Inclusion criteria were: aged 18 to 65 years and a current diagnosis of a major depressive disorder, single episode or recurrent without psychotic symptoms, confirmed by clinical interview and a PHQ-9 score > 8 (Manea et al., 2012; Negeri et al., 2021). Exclusion criteria were acute suicidality, pregnancy or breastfeeding, current neurological or internal diseases, current acute substance abuse (e.g., drug use or excessive alcohol and nicotine use), and a life-time history of psychotic (ICD-10 F20-F29), bipolar (ICD-10 F31) or dissociative (ICD-10 F44) disorder. Patients who fulfilled the inclusion criteria were invited for a brief online meeting through the clinical tool *arztkonsulation.de*. The meeting included a clinical interview conducted by trained psychology students to verify the MDD diagnosis and a screening for all in- and exclusion criteria.

Patients who gave their written informed consent to participate and fulfilled the inclusion criteria were included in the intervention study. They received the digital smartwatch-based intervention with feedback on their own data and filled the PHQ-9 at the end of the study period. The data of the intervention study was compared with a dataset of patients that participated in a previous version of the study (earlier amendment without an intervention). The dataset included 35 patients with the same in- and exclusion criteria as the intervention group from this cohort, comprising two PHQ-9 assessments, one at baseline and one after two to four months.

### Design and Procedures

A total of 86 patients were recruited and invited to participate in the study. Of these, 45 missed their clinical interview or withdrew from the study while 13 dropped out after the interview shortly before receiving the first intervention. Of the 18 remaining patients, one was excluded from the analyses since the PHQ-9 baseline assessment was missing. For six patients missing post-intervention PHQ-9 scores (after 12 weeks) were imputed based on the last observation carried forward from the intermediate PHQ-9 assessments after 8 weeks. Therefore, a total number of 17 patients in the intervention group were analyzed. The retrospective control group included PHQ-9 data (two assessments within a two to four months interval) of 35 patients that took part in a previous version of the SMART study with no intervention. These patients received an offer for the intervention after the study. Thus, in total, data from 52 patients was included in the final statistical analyses, specifically focusing on the initial clinical effects (PHQ-9).

### Feedback and Intervention

Patients who were enrolled in the SMART study and underwent the intervention program (**Table 1**) received weekly reports that included feedback about their progress concerning various smartwatch-based data and their digital self-help interventions. The feedback reports were based on patients’ behavioral data and their responses to the self-ratings (e.g., mood, tension, concentration, alcohol consumption, sleep quality since the beginning of the study, number and quality of social contacts). Patients received standardized weekly psychoeducation and exercises based on evidence-based interventions. These included behavioral activation (e.g., Keijsers et al., 2017) and physical activity exercises (e.g., planning pleasant activities, one of which had to be sports or a more intense activity), psychoeducation about automatic thoughts, social exercises (e.g., expanding and strengthening the social network), as well as cognitive-behavioral exercises for insomnia (i.e., psychoeducation about sleep, sleep hygiene, stimulus control – Vedaa et al., 2020; Lancee et al., 2012, 2013).

**Table 1.**
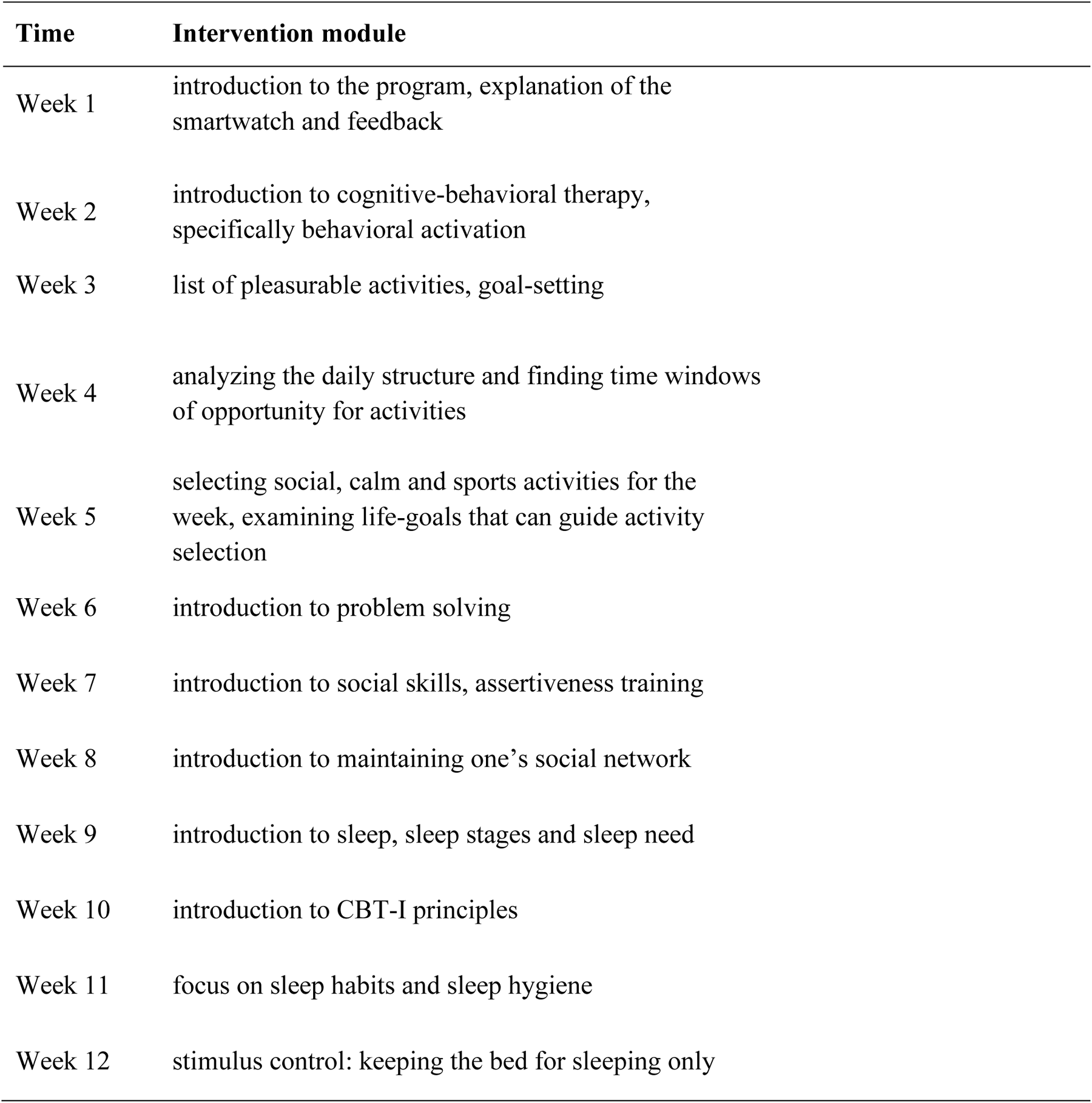
Smartwatch-based intervention program across the 12 weeks.

### Concomitant care

In the intervention group 11 out of the total 17 patients reported to be in treatment, whereas 20 out of the total 35 patients in the control group reported to be in treatment. A chi-square test showed that these proportions were not statistically different, chi-square (1) = 0.27, *p* = 0.602. The treatment was most frequently provided by a psychiatrist or psychotherapist (7 out of 11 in the intervention group, 14 out of 20 in the control group).

## Measures

The SMART study included a clinical interview, as well as repeated measures through questionnaires, smartwatch-based ratings (for feedback purposes) and behavioral parameters. Throughout the study, participants were instructed to wear their smartwatch, which was either the Samsung Frontiers S3 or the Samsung Galaxy Watch, for the entire day.

### Smartwatch EMA

EMA (self-ratings) were carried out throughout the entire intervention period on the smartwatch, beginning after patients were enrolled in the study. It was primarily used for feedback purposes and involved three prompts per day, which was signal-contingent, meaning that patients received automatic prompts from their smartwatch device. We divided a day into three phases (morning: 7:00 AM - 11:00 AM, midday: 11:00 AM - 2:00 PM, and evening: 7:00 PM - 11:00 PM). Then, in each phase, a signal from the smartwatch was emitted and patients had time to complete their self-report until the next time window began. The EMA included 7 different items measuring momentary levels of mood, tension, concentration, sleep quality, alcohol consumption, number of social contacts and number of positive social contacts.

### Questionnaire Assessments

During the first week of the study (t0), participants filled out the main clinical readout of the present study, namely the PHQ-9. The PHQ-9 assesses symptoms of depression of the previous two weeks and meta-analyses have demonstrated its adequate psychometric properties (Gilbody et al., 2007; Kroenke et al., 2001). After the inital PHQ-9 assessment, it was again filled out 8 (t1) and 12 weeks (t2) later.

### Statistical Analyses

To assess the main outcome of the present study—the change in PHQ-9 values from the beginning to the end of the 12-week intervention program in the intervention group compared to those from the control group—we conducted a two-by-two repeated measures analysis of variance (rmANOVA) in JASP 0.18.3 (www.jasp-stats.org). The analysis included group (two levels: intervention and control) as the between-subjects factor, and time (two levels: pre- and post-intervention PHQ-9 values) as the within-subjects factor. The main outcome effect was the group × time interaction. In addition, we performed post-hoc comparative analyses with independent t-tests for both the pre-intervention and post-intervention timepoints. For all analyses the significance level was set at 0.05. All depressed patients, who had aPHQ-9 baseline-value were part of the intent-to-treat analyses, with last-observations carried forward (i.e., for 6 patients PHQ-9 scores at 12 weeks [t2] were missing for which we conservatively used the respective PHQ-9 scores at 8 weeks [t1]). One of the 18 patients from the intervention group had to be excluded as the baseline PHQ-9 score (t0) was missing. We analyzed all patients regardless of exercise performance or increases in physical activity.

## Results

### Demographics of the patients

In total, data from 52 patients was analyzed of which 17 patients were part of the intervention group (age range: 23-54, age *M* = 34.8, *SD* = 9.1, 82% female) and 35 were part of the control group (age range: 19-62, age *M* = 34.7 *SD* = 13.1, 70% female). The groups did not differ in terms of age *t*(50) = -0.02, *p* = 0.982, or depressive symptom severity: *M* = 11.4, *SD* = 4.0 for the intervention group and *M* = 12.6, *SD* = 3.7 for the control group, *t*(50) = 1.04, *p* = 0.303.

### Intervention outcomes

The results of the main analysis are shown in **Figure 1**. At the beginning of the study (t0) both groups, intervention group (*M* = 11.4, *SD* = 4.0) and non-intervention group (*M* = 12.6, *SD* = 3.7) did not significantly differ in their PHQ-9 scores *t*(50) = 1.04, *p* = 0.303, suggesting that the depressive symptom severity was similar before administering the intervention. The group × time interaction was significant *F*(1, 50) = 7.21, *p* = 0.010, indicating that the change in depression severity from pre- to post measurement was dependent on the intervention. There was also a main effect of group *F*(1, 50) = 7.09, *p* = 0.010 but no main effect of time *F*(1, 50) = 0.02, *p* = 0.866. After the intervention, the intervention group had a significantly lower PHQ-9 score (*M* = 9.8, *SD* = 3.5) compared to the non-intervention group (*M* = 13.9, *SD* = 4.1), *t*(50) = 3.55, *p* < 0.001.

**Figure 1.**
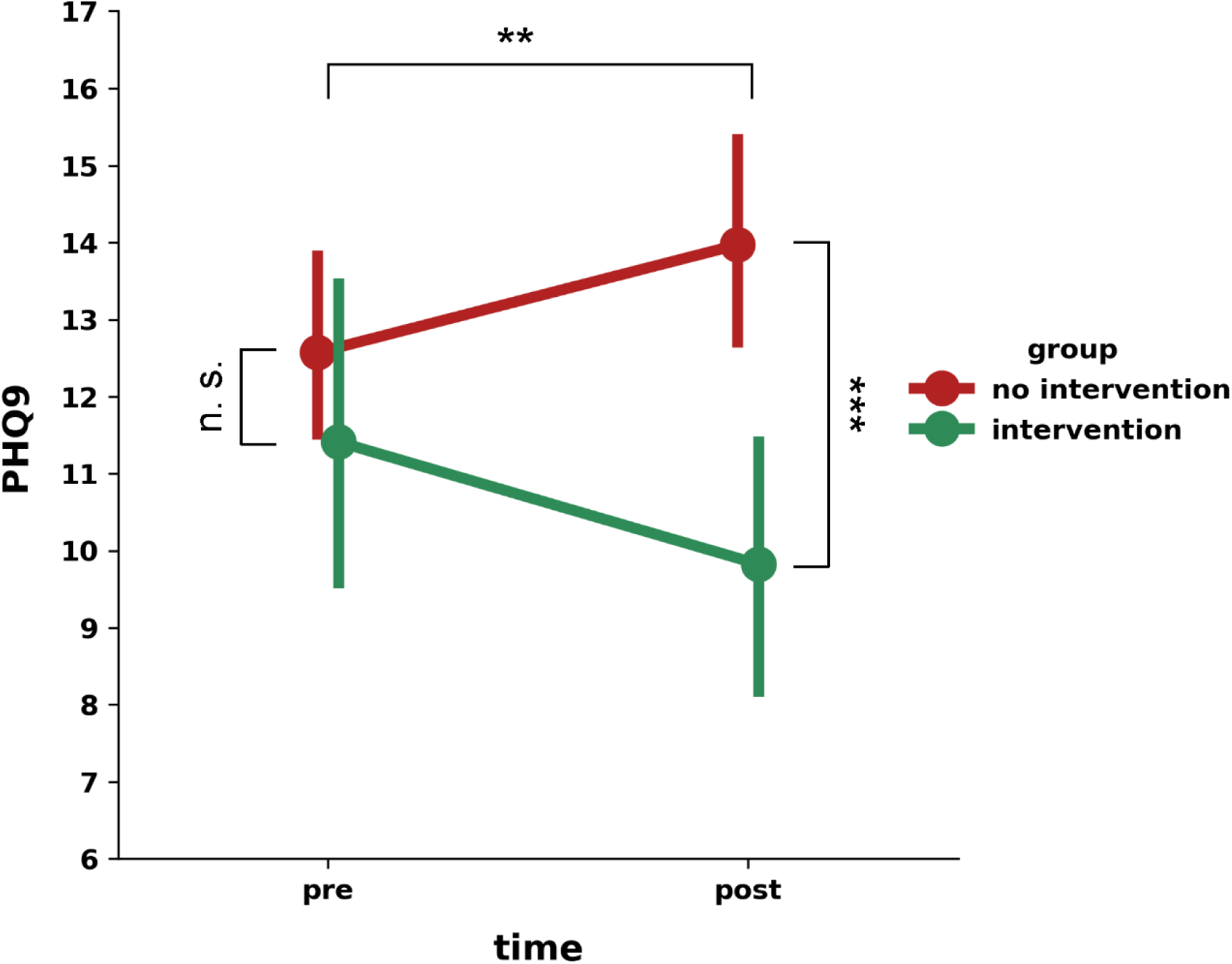
Change in Depression Severity (PHQ-9) from pre to post in the Intervention and Control Group. *Note*. Comparison of PHQ-9 scores of the intervention and retrospective control group. The plot shows the mean values including error bars for the pre- and post-intervention phase for both groups, i.e., intervention and non-intervention group. Significance level is indicated as follows: ***p < 0.001, **p < 0.01, n.s. = not significant.

## Discussion

This study aimed to examine the initial effects of a smartwatch-based intervention program for patients with mild to moderate depression. We collected data from two groups undergoing therapy: one group (the intervention group) received the smartwatch-based intervention, whereas a retrospectively determined control group did not. To evaluate the benefits of the smartwatch intervention, we compared PHQ-9 scores of both groups at baseline and after 12 weeks of intervention versus waiting time. We found that patients that underwent our smartwatch-supported intervention program showed an improvement in depressive symptomatology. These findings are consistent with two previous digital intervention studies (Aguilar-Latorre et al., 2022, 2023) that demonstrated superior intervention effects of a lifestyle modification program, combining the use of smartwatches with weekly group therapy sessions. The comparison of this program with treatment as usual (i.e., medical care by general practitioner) for depression indicated positive effects for the patients in the lifestyle modification program. Our findings extend these findings by showing that an automated, purely digital intervention that integrates objective and subjective feedback from smartwatches and provides reminders shows similar promising effects.

## Limitations

This pilot study has several limitations. First, the sample size of this effect size estimation pilot-study was small, which limits the statistical power of the results. Second, we compared the effects of the smartwatch-based intervention to a retrospectively determined control group that received no intervention. Consequently, future research is needed to validate the results with a prospective randomized controlled trial. In such a trial, it would be informative to compare the effects of such a smartwatch-based intervention with a smartphone-based intervention, to evaluate the potential effects of saliency in its usage and more accurate feedback on physical activity. The combination of having a device continuously in sight with prompts that remind the patient to perform actions, while being aware that activity levels are monitored and feedback is provided, might be motivating and enhancing compliance. This seems to be a secondary feature for common digital therapies, which typically focus on contents. However, the saliency, which smartwatches offer, could be essential given the low adherence rates to other digital therapies. Moreover, it allows the closing of an activity-related feedback loop between intervention (planning activities) and monitoring (what was actually performed). In this initial study, our activity-related feedback only comprised step-counts but this could readily be extended to additional feedback such as GPS pattern diversity, sleep efficiency or activity intensity, timing and variability. Such feedback loops could form the basis for more stratified and eventually individualized exercises and psychoeducative materials. This would further show the relevance of incorporating objective data to increase physical activity levels and conduct behavioral activation beyond subjective impressions of activity.

## Conclusion

This study demonstrates the feasibility and efficacy of a smartwatch-supported digital intervention for reducing depression severity. While in this study the smartwatch device was mainly used for subjective ratings and physical activity, the potential of smartwatches in tracking depression-related information is not limited to these variables. Instead, further variables associated with depression including sleep efficiency, light exposure and location data can be continuously monitored (Sequeira et al., 2019; Triantafyllidis, 2024). The easy-to-use interface and adaptable module structure of smartwatches make them a promising tool for enhancing psychotherapeutic treatment, outsourcing specific modules or parts thereof to the digital domain. Additionally, the use of smartwatches can address the incorporation and monitoring of physical activity exercises into common treatment schemes, offering a practical solution for healthcare providers being limited to in-house healthcare facility treatment. These advantages of smartwatches are not necessarily restricted to depression, but could be extended to the treatment and prevention of other mental disorders and chronic diseases.

## Data Availability

All data produced in the present work are contained in the manuscript.

## Acknowledgments

The authors would like to thank Florian Binder for his technical support.

## CRediT

Conceptualization: JT, MF, VS; Methodology: JT, AB, VS; Recruitment and data collection: JT, AEB, MF, DS, DP, JG, JS, RB, RD; Data curation: JT, AB, AEB, DS; Formal analysis: JT, AB, AEB, VS; Writing-original draft: JT, AB, VS; Review of first draft: all; Supervision: VS; Project administration: JT.

